# Modeling Longitudinal Perception-Based Recovery Through the Use of Standard Stroke Recovery Metrics

**DOI:** 10.1101/2025.06.30.25330599

**Authors:** Yash Akkara, Kevin V. Nguyen, Michael Lemonick, Ryan Afreen, Christopher P. Kellner

## Abstract

**Background and Purpose:** Stroke is a leading cause of death and disability worldwide. Among the various domains affected by stroke, perceptual deficits are particularly under-recognized despite their significant impact on a patient’s quality of life. To address this gap, we conducted a systematic review to evaluate longitudinal changes in perception-recovery following stroke and developed an assessment battery using standardized stroke metrics to assess perceptual function.

**Methods:** Following PRISMA (Preferred Reporting Items for Systematic Reviews and Meta-Analyses) guidelines, studies using standardized stroke assessment metrics in patients with either ischemic or hemorrhagic stroke were screened using PubMed, MEDLINE, and Embase. Studies that included participants with prior neurological disability or new interventional treatment (including conservative, medical, and surgical management) were excluded.

**Results:** A total of 120 studies with 71,179 patients were included. Neurological recovery showed a two-phase recovery pattern, with the most rapid improvements in the first year followed by slower, fluctuating changes. In contrast, tools measuring motor ability, cognition, balance, and perceived health revealed a steady recovery concentrated within the first year, with a plateau afterward. Mood improved gradually over time, and quality of life showed consistent gains. We also identified specific items within each metric that correspond to individual subdomains, based on primary validation data and existing stroke literature, to create a perception-based assessment battery.

**Conclusions:** These patterns suggest that perceptual, cognitive, and functional recovery after stroke may continue well beyond the early phase, highlighting how scores evolve over time. By modeling longitudinal changes, we aim to support clinicians in understanding perception-based recovery and encourage further research into its prognostic value.

## Introduction

Stroke is the second leading cause of mortality worldwide and presents a growing global health challenge due to its increasing prevalence, long-term disability, and economic impact.^1–3^ Furthermore, effective stroke recovery is a vital component of patient care, aiming to restore function, enhance quality of life (QoL), and reduce long-term disability, both within and beyond the clinical setting. Since stroke can affect various cognitive domains such as language, motor control, and memory, clearly categorizing these domains is essential to guide targeted interventions.^4–6^ However, current literature regarding stroke recovery categorization is limited, primarily due to the heterogeneity of stroke and stroke recovery.^7^ To address this deficit within the literature, we conducted a previous systematic review identifying seven distinct domains of stroke recovery–perception, physical and motor function, speech and language, cognition, activities of daily living, quality of life, and social interaction–to develop a standard stroke recovery metric that could holistically measure longitudinal recovery across these various domains.^8^

This paper focuses on the domain of perception, which is defined as the brain’s ability to interpret sensory input such as sight, sound, touch, and smell.^9^ Stroke-related perceptual deficits can persist long-term and significantly impair an individual’s QoL.^9–11^ Within the domain of perception, we identified 15 key subdomains that represent a wide spectrum of important sensory, cognitive, and motor functions.^8^ These subdomains are: level of consciousness, vision, pain, neglect, orientation, vitality, static balance, dynamic balance, discomfort, touch, proprioception, sleep, appetite, restlessness, and sensitivity to stimuli. We also identified other less frequently measured, but crucial, subdomains of perception, such as appetite, restlessness, sensitivity to stimuli, and allodynia (Table 1).^8^

**Table 1.**
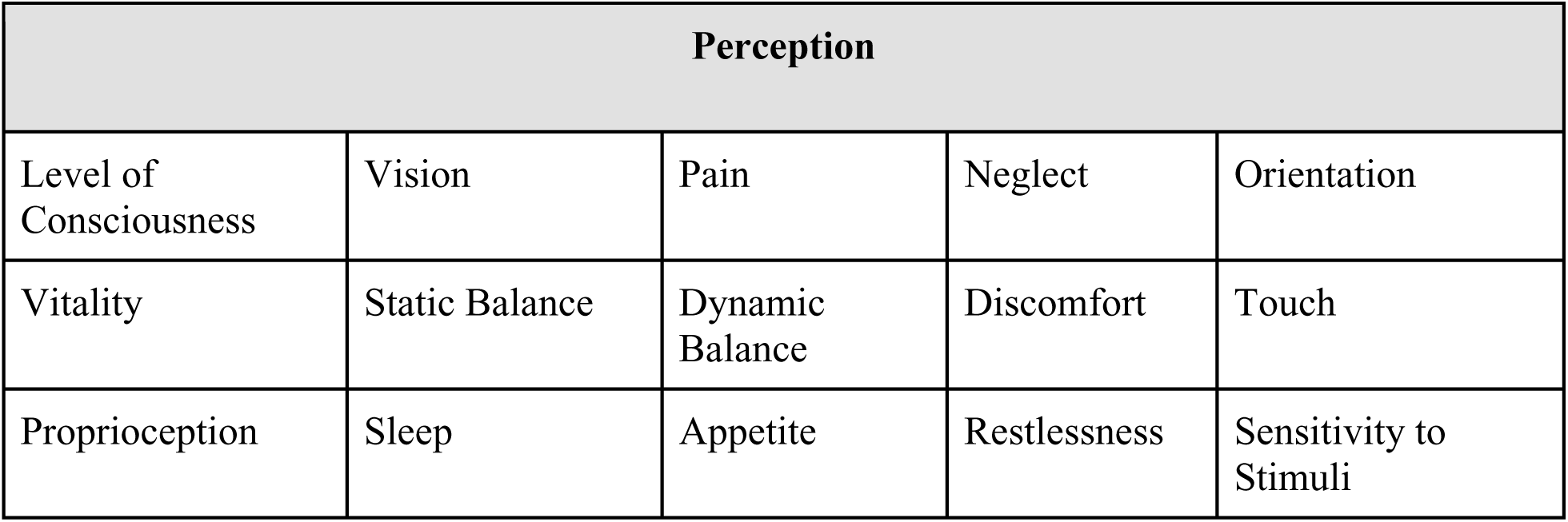
Subdomains of Perception.

Since 2004, there has been a growing focus in the literature on assessing perception after stroke.^8^ However, despite its increasing utilization, perceptual deficits are still under-detected in many patients.^9,12^ Additionally, current guidelines for assessing perception post-stroke lack clarity and consistency, creating challenges in standardizing care and tailoring rehabilitation approaches.^9,13^ This is a common concern among healthcare professionals because perception encompasses a complex coordination of systems in order to function, leading to significant variability in how guidelines are interpreted and care is delivered.^9,14^ To address this gap, we analyze how different stroke assessment metrics change longitudinally and introduce a novel assessment tool that clinicians can use to measure perception recovery more systematically.

## Methods

The methodology employed in this study is based on the protocol established in our prior research and serves to build on the domain-specific metrics we developed by characterizing their expected trajectory in a standard stroke population, alongside merging their elements to develop a combined, domain-specific assessment battery.^8^ Key parameters, particularly with regard to screening, were retained to ensure consistency. As such, similar to our original study, a structured literature search was conducted in accordance with the systematic review guidelines outlined by the Preferred Reporting Items for Systematic Review and Meta-Analysis (PRISMA) checklist. Articles from January 2004 to March 2025 were searched using PubMed, MEDLINE, and Embase. Only articles in English were included. The keywords “Stroke Recovery OR Stroke Rehabilitation AND Domain AND Measure OR Metric OR Test OR Assessment OR Questionnaire” were used in MeSH and text-word format.

However, to focus on the domain of perception and the metrics we identified, we altered our inclusion criteria to ensure studies provided longitudinal measurements of scores in the NIH Stroke Scale (NIHSS), EuroQol-5D (EQ-5D), Fugl-Meyer Assessment-Upper Extremity (FMA-UE), Mini-Mental State Examination (MMSE), Berg Balance Scale (BBS), Geriatric Depression Scale (GDS), and WHO Quality of Life-BREF (WHOQOL-BREF). Moreover, it was required that all data regarding longitudinal changes in the aforementioned metrics was validated in a standard stroke population undergoing the standard-of-care. As such, studies with participants with prior neurological disability or new interventional treatment (including conservative, medical, and surgical management) were excluded. Inclusion was limited to experimental and observational cohort studies in English, consisting of patients with either ischemic or hemorrhagic stroke. Due to the differences in pathophysiology and treatment, alongside variance in the metrics used to assess recovery, patients with subarachnoid hemorrhage (SAH) were excluded from this analysis. All studies were required to have a sample size of ≥20 participants, who were all ≥18 years of age. No articles were excluded based on the duration following the diagnosis of stroke. All studies were required to collect longitudinal recovery data, with a minimum follow-up of 90 days following the initial/baseline measurement. In cases where a study contained a partial population that fit the inclusion criteria with sufficient details regarding their demographic details and follow-up outcomes, only subjects/treatments that were relevant to our inclusion criteria were included.

Studies from the database search were entered into the systematic review software Covidence. Three reviewers independently completed the initial abstract and title screening, followed by the full-text screening in accordance with the inclusion and exclusion criteria. Any conflicts in decisions were discussed until a consensus was reached with the assistance of a senior author. Data extraction was then completed based on the following set of outcomes:

*Author, study name, study design, country, sample size, male:female ratio, population age (mean, median, range), mechanism of stroke, metric(s) used, mean/median measurements of the metric in a standard population (baseline, up to 48 months and beyond), number of patients constituting each measurement, dropout rate, and follow-up*.

The Newcastle-Ottawa Scale (NOS) was used for observational studies. The Cochrane Risk of Bias 2.0 tool was used for interventional studies. Quality assessment(s) were conducted independently by all reviewers (Y.A., R.A., M.L., K.N.). Studies using the NOS were scored from 0 to 9, while studies using the Cochrane Risk of Bias 2.0 tool were scored based on a low, moderate, or high level of bias. Discrepancies in scoring were first discussed between authors and were escalated to a senior author (C.K.) in persistent conflicts, who also evaluated the study for bias to arrive at a judgment.

Following the pooling of all participant demographic and longitudinal data, the Shapiro– Wilk test was used to establish normality, with the two-tailed T-test and Spearman’s R used to test for differences between participant demographic factors across the metrics. Data for all metrics at their respective time points were pooled across studies using a random-effects model and weighed according to the number of participants constituting each measurement by study. Following pooling at each time point, a non-linear regression was performed using iterative processing to identify the best suitable model for the data. 95% confidence interval bands were plotted alongside the regression fit, with each data point’s coefficient of variance (CV) graphed alongside. To develop a combined assessment battery for the perception domain, the initial source of each metric was consulted to identify the item on each questionnaire corresponding to the specific subdomains. Multiple metrics testing the same subdomain of interest were tested for their predictive value to each other, and tertiary stroke-specific metrics were used to identify the more appropriate test for inclusion. When insufficient data was present to directly compare multiple metrics, the more frequently validated metric in the literature was chosen for its specific subdomain. Data was compiled and analyzed using GraphPad Prism Version 10.1.1 for Mac (GraphPad, San Diego, CA, USA). Parametric variables are presented as mean (± standard deviation). Nonparametric datasets are presented as a median (interquartile range). p < 0.05 was considered statistically significant. This review has been registered with the International Prospective Register of Systematic Reviews (PROSPERO). Registration ID: CRD42024551753.

## Results

A total of 120 studies were included in the final analysis, of which 69, 11, 18, 9, 5, 4, and 4 studies used the NIHSS, EQ-5D, FMA, MMSE, BBS, GDS, and WHOQOL-BREF, respectively. The majority of studies were from the WHO European region, followed by the Americas, and the Western Pacific and South-East Asia regions, respectively. Most studies were from populations located within high-income countries, followed by some studies from upper-middle and lower-middle income countries, respectively. These findings are described in Figure 1.

**Figure 1.**
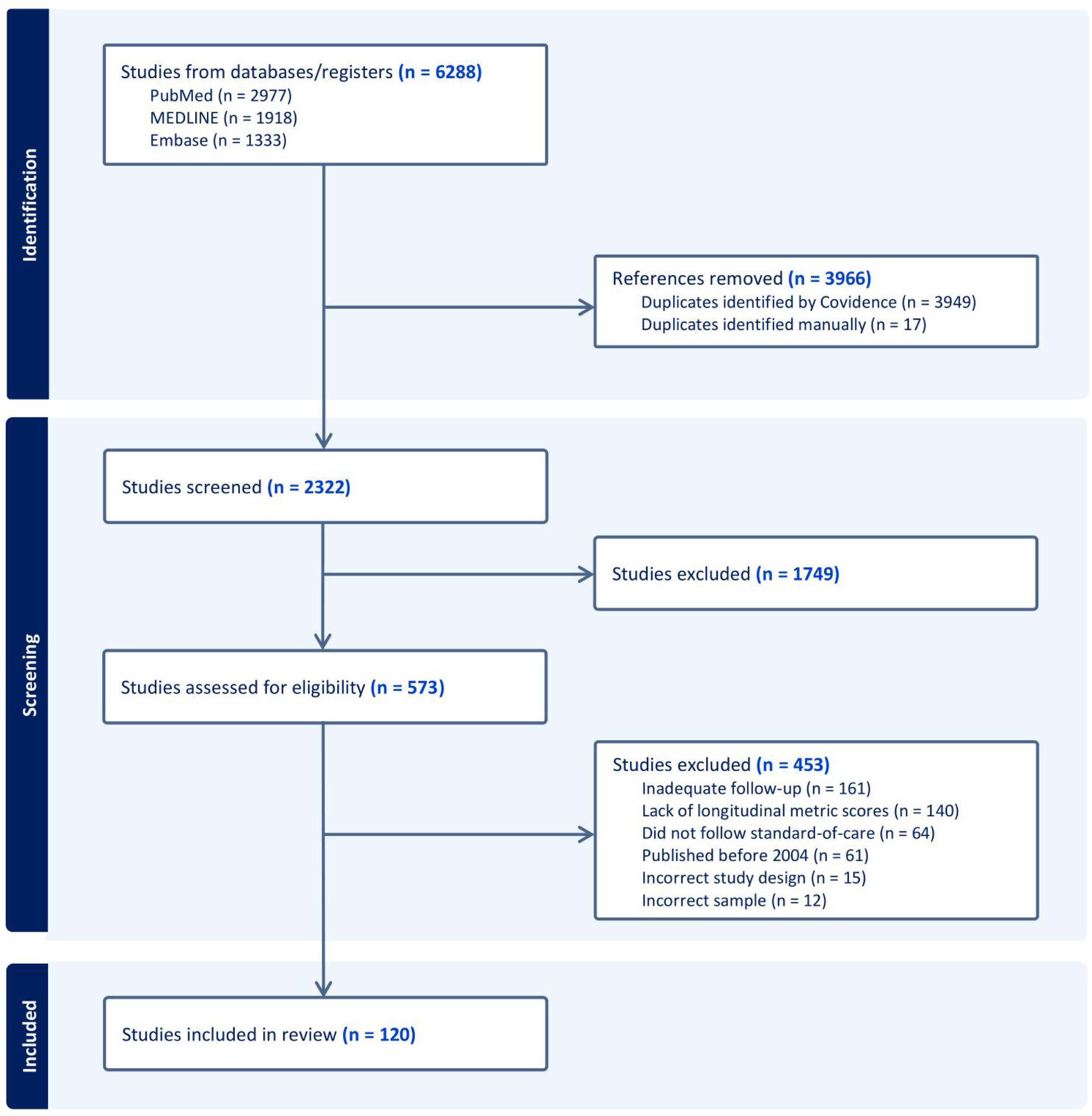
PRISMA diagram outlining the stages of screening and inclusion of studies.

A total of 71,179 participants were included across all studies. Of these, 45123, 19598, 1986, 2143, 509, 940, and 880 participants were tested using the NIHSS, EQ-5D, FMA, MMSE, BBS, GDS, and WHOQOL-BREF, respectively. All participants formed a standard stroke population, with no additional or experimental management provided. Participant demographic characteristics are described in Table 2.

**Table 2.**
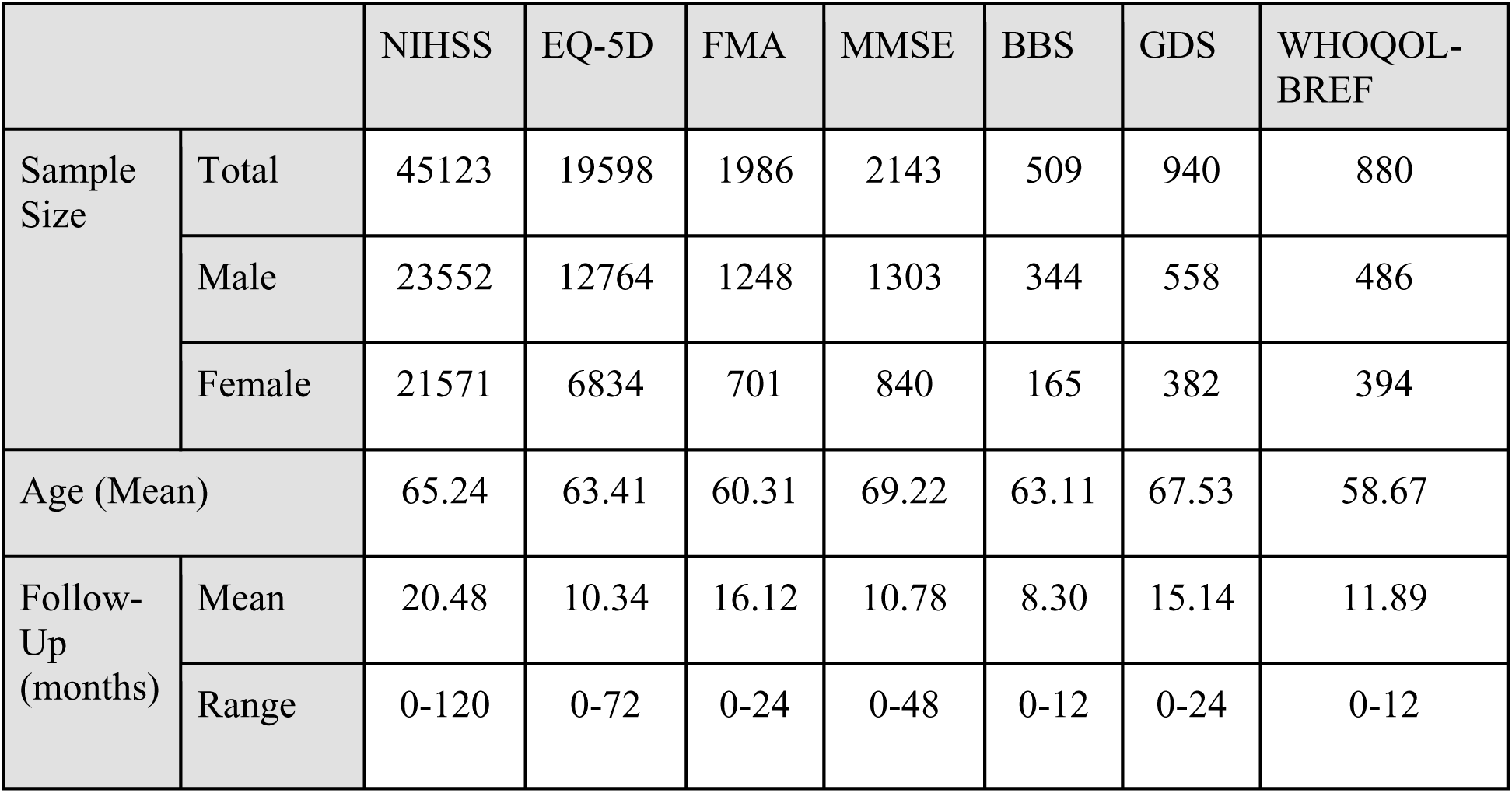
Demographic Characteristics of Participants.

|  |  | NIHSS | EQ-5D | FMA | MMSE | BBS | GDS | WHOQOL-BREF |
| --- | --- | --- | --- | --- | --- | --- | --- | --- |
| Sample Size | Total | 45123 | 19598 | 1986 | 2143 | 509 | 940 | 880 |
|  | Male | 23552 | 12764 | 1248 | 1303 | 344 | 558 | 486 |
|  | Female | 21571 | 6834 | 701 | 840 | 165 | 382 | 394 |
| Age (Mean) |  | 65.24 | 63.41 | 60.31 | 69.22 | 63.11 | 67.53 | 58.67 |
| Follow-Up (months) | Mean | 20.48 | 10.34 | 16.12 | 10.78 | 8.30 | 15.14 | 11.89 |
|  | Range | 0-120 | 0-72 | 0-24 | 0-48 | 0-12 | 0-24 | 0-12 |

Following the modeling of all 7 metrics, non-linear curve regressions were performed based on iterative best-fit modeling. The NIHSS model (Figure 2A) was best predicted by a mixed model, using both exponentially declining and sinusoidal regressions (R^2^ = 0.9721, R^2^_adj_ = 0.9582, RMSE = 0.6002). Initially, mean NIHSS scores experienced exponential declines, with a relative plateau up to 4 weeks, followed by a sharp decline in scores up to 1 year. Following the first year, however, the data was modeled by a sinusoidal relationship with a low relative amplitude, oscillating roughly every 2 years.

**Figure 2.**
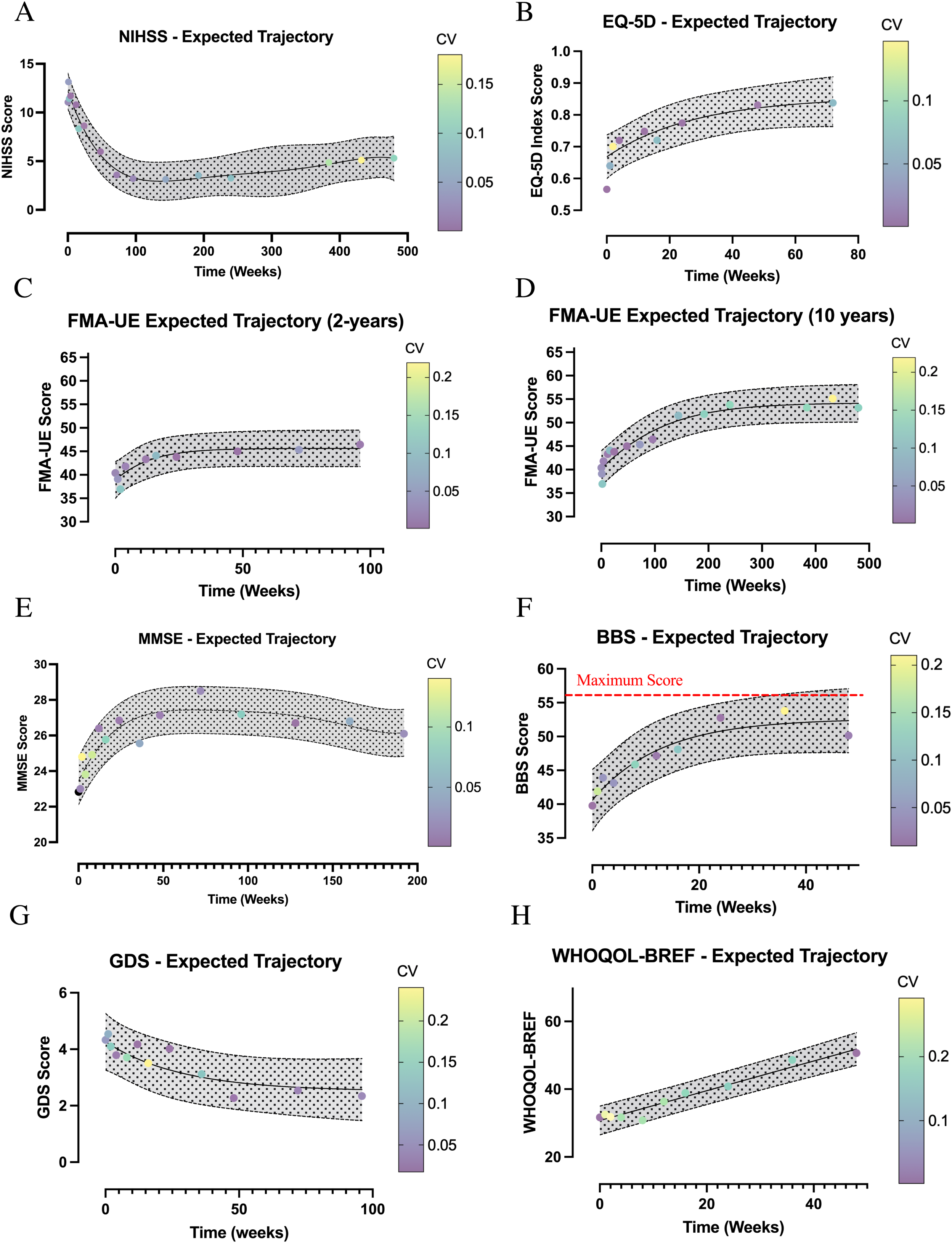
Trajectory of Mean Scores (with 95% CI) of the NIHSS (A), EQ-5D (B), FMA-UE up to 2 years (C), FMA-UE up to 10-years (D), MMSE (E), BBS (F), GDS (G), and WHOQOL-BREF (H). Across all panels, the solid lines represent the modeled mean scores at each follow-up time point, while the shaded regions denote 95% confidence intervals. The x-axis displays recovery time in weeks, while the y-axis shows the mean numerical score for each metric. CV denotes the coefficient of variation. Dot color reflects the magnitude of variability at each time point. In Panel F, the maximum score for the Berg Balance Scale (BBS) is 56, indicated by a red dotted line.

The EQ-5D was best predicted using a logarithmic model (R^2^ = 0.9053, R^2^_adj_ = 0.8737, RMSE = 0.02091), displaying significant growth up to 3 months following stroke, after which it experienced plateaus leading up to 1-2 years post-stroke (Figure 2B).

For the FMA-UE (Figure 2C), we identified studies with data up to 2 years, with the model displaying a significant logarithmic trajectory, increasing within the first 3 months following stroke, and experiencing a significant plateau up to 2 years (R^2^ = 0.8321, R^2^_adj_ = 0.7841, RMSE = 1.235). We also achieved a low level of variance at these time points, owing to relatively high usage of the metric. We also performed an additional analysis for certain studies reporting data up to 10-years (Figure 2D), finding a relative but gradual increase in scores continued to be observed at a lower level of confidence, maintaining reasonable logarithmic growth up to 4 years post-stroke (R2 = 0.9330, R2adj = 0.9227, RMSE = 1.498).

The MMSE (Figure 2E), was best modeled by logarithmic and polynomial (cubic) growth (R^2^ = 0.8148, R^2^_adj_ = 0.8148, RMSE = 0.6741). Up to 2 years, the MMSE was best predicted by a logarithmic growth equation, reaching a plateau following 1 year of a general increase in mean scores. Following the 2-year mark, the MMSE was best modeled with a cubic polynomial equation. While the polynomial model results from the iterative processing and is itself unlikely to represent the real relationship (especially at longer follow-ups), the model showed a gradual decline in scores over time following the plateau, accompanied by a partial, undefined cyclability.

The trajectory of mean scores in the BBS (Figure 2F) was best modeled by exclusively logarithmic growth (R^2^ = 0.9073, R^2^_adj_ = 0.8808, RMSE = 1.411). The model shows a relative improvement in scores up to 3 months post-stroke, followed by a plateau up to 12 months. Due to the scarcity of studies and iterative modeling in the BBS, we were unable to achieve a low level of inter-study variance, with insufficient data for modeling beyond 1 year.

The GDS (Figure 2G) was modeled exclusively using negative logarithmic growth (R^2^ = 0.7592, R^2^_adj_ = 0.7056, RMSE = 0.3503). The model highlighted significant reductions in overall scores up to ∼1 year, following which a plateau was observed, with mean scores only decreasing marginally up to 2 years and beyond.

Finally, the WHOQOL-BREF (Figure 2H) was best modeled by linear growth (R^2^ = 0.9532, R^2^_adj_ = 0.9473, RMSE = 1.584). Although some general fluctuations were observed over time in the mean scores of participants, scores were found to increase at a generally stable rate. Similar to the BBS, due to the scarcity of usage of the WHOQOL-BREF, we were unable to successfully model growth beyond 1-year post-stroke.

In an effort to reduce levels of redundancy by testing the subdomains within the overall domain of perception, we identified the specific item(s) in each metric corresponding to the specific subdomain being tested, based on primary data from metric validation, and through the larger body of secondary stroke literature. While testing of the complete metric may still be required if scoring is required, this categorization enables clinicians to identify the specific purpose of relevant components of metrics to testing perception, and hence can allow the isolation and repeated testing of these items to characterize subdomain-specific recovery in a longitudinal manner, when needed. This is described in Supplementary Table 1.

## Discussion

Stroke is a leading cause of disability and death worldwide, often resulting in significant impairments in both motor and cognitive functions. Among the various consequences of stroke, perceptual deficits are particularly challenging to address because perception encompasses a variety of complex cognitive processes, like spatial awareness, attention, and the ability to interpret sensory information, making it difficult to standardize rehabilitation efforts. The present study provides a comprehensive analysis of how general mean scores of perception, measured by the NIHSS, EQ-5D, FMA, MMSE, BBS, GDS, and WHOQOL-BREF, change over time in stroke populations. Additionally, this study identifies the parts of each metric that correspond to the subdomains of perception, allowing clinicians to assess and monitor individual components of perception on a more systematic level.

The results of the present study utilized best-fit non-linear regression models to outline the recovery trajectories of several key outcome metrics commonly used in stroke recovery research. It is important to note that, within our analysis, we show the trajectory of mean scores of perception, as measured by the NIHSS, EQ-5D, FMA, MMSE, BBS, GDS, and WHOQOL-BREF, for a standard stroke population. These mean scores reflect the general trends of perceptual recovery over time and the 95% confidence interval within which the mean score of the stroke population is likely to fall at each time point. Therefore, although this analysis provides valuable insight into the average recovery pattern of perception, which can help manage clinical expectations and rehabilitation strategies, these results are not meant to be specific cut-offs for what patients should be scoring at each time point in their recovery. For any given time point, the 95% confidence interval of expected scores for individual patients could be much broader because of the variability in stroke recovery across different individuals.^9,13^ Nevertheless, by examining the changes in mean scores, as assessed by the aforementioned metrics, we uncovered distinct patterns of improvement, plateau, and fluctuation in perception.

Our analyses revealed that recovery following stroke is not a uniform process, but one characterized by different recovery phases. In the NIHSS, mean perception scores were best modeled using a combination of exponential decline and sinusoidal regression. Our findings show that immediately following a stroke, there is a rapid decline in NIHSS scores, signifying an improvement in function, that extends to the 64-week mark. This is consistent with previous studies showing that the first 12 weeks following a stroke is the most crucial period for recovery, although recovery can extend beyond 52 weeks post-stroke.^15,16^ Around the 128- to 512-week mark, there was an increase in mean NIHSS score, which was modeled by a sinusoidal relationship. This later increase in scores may, in part, reflect a deterioration or fluctuation in neurological status that may be attributed to a variety of factors such as cognitive declines related to aging, comorbidities, recurrent cerebrovascular events, or other long-term complications like depression.^17–19^ Furthermore, at these later time points, our CV was much higher than at earlier time points, suggesting greater individual differences in the long-term recovery of perception and the need for continued long-term assessment of perception recovery following stroke.

The FMA-UE scale is a widely used metric to assess motor impairments following a stroke. The FMA-UE consists of 33 items, and each item is scored on a scale of 0-2 (0 = cannot perform, 1 = performs partially, and 2 = performs fully), resulting in a total of 66 possible points.^20^ Motor recovery, as measured by the FMA-UE, revealed two distinct sub-trajectories: one within the first two years post-stroke, and a second extending up to ten years. Up to two years, FMA-UE scores followed a significant logarithmic trajectory, with most improvements occurring within the first three months, followed by a plateau through the two-year mark. This pattern suggests that motor recovery improves drastically within the first few months after a stroke, with a gradual slowing in improvement thereafter.^21^ Previous studies have also described an increased synaptic plasticity period of 1-3 months after a stroke in which rehabilitation efforts are crucial to improving post-stroke recovery.^22,23^ Interestingly, we also found that after 2 years, improvements in motor function still continued, albeit, at a slower rate compared to the first 2 years, through the 10-year mark. However, these long-term gains were accompanied by increasing coefficient of variation (CV) values, indicating greater variability in motor recovery outcomes across individuals.^24^ Although, long-term measurements including the FMA-UE is sparse in the literature, Ingwersen et al., previously reported FMA scores could improve by about 4 points per year in linear random coefficients models.^25^ These results suggest that motor recovery, as measured by the FMA-UE, has a potentially long period for improvement in patients who have had a stroke. Thus, it is important for clinicians to recognize that meaningful gains can occur well beyond the acute and subacute phases, and to incorporate long-term rehabilitation planning into patient care. Future research should also examine the long-term recovery trajectory of the lower extremities using the FMA-LE.

The EQ-5D is commonly used to assess health-related QoL in stroke patients and measures five dimensions: mobility, self-care, usual activities, pain/discomfort, and anxiety/depression.^26^ Items are scored based on the level of severity (Levels 1-3) experienced by the patient.^26^ Mean EQ-5D scores were best modeled by a logarithmic curve, which reflected the significant growth in health-related quality of life up to 3 months following stroke. This was followed by a slight plateau extending up to 1-2 years post-stroke. This pattern represents a consistent trend in health-related QoL recovery, where the most substantial improvements occur early on, and the rate of change diminishes as health recovery reaches its peak.^27,28^ Interestingly, previous studies have suggested that there may be sex differences in health-related QoL following stroke, in which women have worse QoL than men up to 12 months after stroke.^29^ This emphasizes the need for the continued assessment of QoL in stroke patients over time, as it suggests that there may be a possible sexual dimorphism in QoL outcomes.

The MMSE is a metric that assesses cognitive status, comprising 11 questions across a variety of cognitive domains. The maximum score is 30 points, and scores of 23 or lower suggest cognitive impairment.^30^ In our analysis, MMSE trajectory, modeled using logarithmic and cubic polynomial growth, points to rapid early improvements in cognitive function that plateau around the 40-week mark. However, there is a slight decline in mean MMSE scores following the initial plateau, suggesting delayed cognitive impairment in some patients following a stroke. This pattern aligns with previous studies reporting a significant increase in 5-year dementia risks in stroke survivors compared to individuals who have never experienced a stroke.^31,32^ In fact, the American Heart Association/American Stroke Association has reported that the incidence of new-onset dementia was 14.2% at 1 year, increasing to 28.3% at 4 years following stroke.^33^ Other factors could also contribute to the decline in mean MMSE scores, such as age-related cognitive declines or potential secondary cerebrovascular accidents.^34,35^ Notably, MMSE scores were lowest around the 200-week mark, and reflect a deterioration in cognitive function.^30^ This cognitive deterioration, as measured by the MMSE, coincides with an increase in the NIHSS trajectory, which also occur close to the 200-week mark. Since the NIHSS encompasses various neurological domains such as level of consciousness, language, and motor function, a worsening in cognitive performance may correlate with or even exacerbate these deficits, ultimately resulting in higher NIHSS scores.^36^ Taken together, our results suggest that the majority of mental health improvement post-stroke, as measured by the MMSE, occurs before the one-year mark, and that, in the years following stroke, delayed cognitive decline can occur. Additionally, these findings underscore the importance of long-term cognitive monitoring in stroke survivors, as initial recovery may mask the emergence of delayed cognitive and neurological decline.

Balance recovery is often measured by the BBS, a common assessment tool that measures multiple dimensions of balance through a series of functional tasks with reliable reproducibility.^37^ The test consists of 14 items that assess daily activity performance, each scored from 0 (unable) to 4 (independent). The total score ranges from 0 to 56, with higher scores indicating better balance.^37^ In the present study, the trend in BBS scores was best described by a logarithmic model. Our results suggest that patients who have had a stroke experience the majority of balance recovery within the first three months post-stroke, followed by a plateau through twelve months.^38,39^ The observed plateau may be influenced by several factors. First, balance is strongly influenced by cognitive function.^40,41^ Since mental awareness and cognitive ability have been shown to improve significantly within the first three to month months post-stroke, this may lead to a stagnation in balance control improvement and the potential for increased fall risk over time.^38,40–42^ Second, age-related musculoskeletal changes, such as decreased muscle strength, joint flexibility, and proprioception, may contribute to stagnation or even slight declines in balance scores.^43,44^ It is worth recognizing that studies using the BBS that met our inclusion criteria were limited, restricting BBS modeling to twelve months. Thus, long-term, continuous balance assessment in stroke patients represents an important area for clinicians to measure to prevent secondary complications following stroke, like falls, loss of mobility, and decreased independence.

Post-stroke depression (PSD) is a common complication, affecting approximately 30% of stroke survivors.^45^ The GDS is a 30-item questionnaire that helps to identify depression in older adults.^46^ Each question is assigned either 1 point (if a patient answers “yes”) or 0 points (if a patient answers “no”), with higher scores correlating with more severe depression.^46^ In the case of depressive symptoms measured by the GDS, our results were modeled exclusively by a negative logarithmic model that revealed significant early improvements up to around one year, with a minimal decline thereafter. These results suggest that most improvements in PSD symptoms occur within the first year, after which recovery plateaus.^45,47^ Additionally, even though a multitude of factors can contribute to the development and persistence of PSD, previous studies have shown that early depressive symptoms following a stroke are significant predictors of PSD.^45^ In fact, among those who were depressed within 3 months of stroke, 53% of patients experienced persistent depression even after one year.^48^ Thus, clinicians need to screen patients for potential depressive symptoms and continue to repeatedly monitor patients long-term, as early identification and treatment of depression can directly improve patient outcomes.^33^

The WHOQOL-BREF measures QoL across four domains: Physical Health, Psychological, Social Relationships, and Environment.^49^ Each item is rated on a 5-point Likert scale in which higher scores indicate a better quality of life.^50^ Outcomes regarding QoL, as measured by the WHOQOL-BREF, were best modeled by a linear growth function with some general fluctuations. Our results suggest that there is a steady increase in an individual’s perception of their QoL following a stroke that extends to the one-year mark.^51,52^ Interestingly, although both the EQ-5D and the WHOQOL-BREF both measure health-related quality of life, the trajectory of the EQ-5D appears to reach a plateau before the WHOQOL-BREF. This may be because while both measure aspects of health-related QoL, they emphasize different dimensions of perception recovery. The EQ-5D is a more concise and objective tool that primarily assesses functional health states, such as mobility, self-care, and pain/discomfort, which may reach a plateau earlier as physical recovery stabilizes.^28^ On the other hand, the WHOQOL-BREF incorporates broader psychosocial factors like emotional well-being, social relationships, and environmental adaptation, which may continue to evolve over time.^53,54^ Additionally, the linear growth expressed by the WHOQOL-BREF scores suggests that even as physical health stabilizes, individuals may still experience improvements in psychological adjustment, social reintegration, and overall life satisfaction.^55,56^ Thus, the subjective nature of the WHOQOL-BREF and an individual’s perception of their life and ability to adjust to their environmental circumstances may contribute to the observed linear growth pattern.

The findings from our study reveal distinct but interconnected patterns of recovery across multiple domains. Across nearly all metrics, an initial phase of logarithmic or exponential recovery was observed, particularly within the first year post-stroke, implicating the role of neuroplasticity and the critical window for rehabilitation and intervention following stroke.^15,16,51^ Additionally, we were able to model the long-term trajectories for some metrics like the NIHSS, FMA-UE, MMSE, and GDS, extending from one to ten years. Thus, these results could provide valuable insight into how stroke recovery evolves over a decade and allow clinicians to gauge how their patients are recovering compared to the rest of the stroke population. For the EQ-5D, BBS, and WHOQOL-BREF, long-term analysis that surpassed a year was unable to be modeled due to the lack of available data in the literature beyond the one-year mark. However, the eventual plateaus of these metrics suggest that perception recovery after one year, as measured by the EQ-5D, BBS, and WHOQOL-BREF, is minimal and emphasizes the importance of early rehabilitation strategies to optimize recovery outcomes. Despite this, it is still important to continue assessing patients beyond the first year to monitor for any subtle, long-term changes that may occur.^57^

In our previous work, we have identified 15 subdomains that make up the complex nature of perception and the metrics that best assess each subdomain.^8^ Perception has been difficult to define and standardize and arises from the fact that perception is not a singular function but rather a combination of individual, but interrelated subdomains, each responsible for specific aspects of sensory and cognitive processing.^9^ As a result, the literature lacks a standardized way to efficiently and effectively measure a patient’s perception following a stroke.^9,58^ To address this issue, we added a combined battery incorporating the seven previously identified stroke metrics to address these subdomains of perception (Supplementary Table 1). This combined metric aims to address the challenges associated with defining and standardizing perception by offering a structured approach that isolates and measures each subdomain individually. In the clinical setting, this allows physicians and other health professionals to focus on specific items of concern across repeated, longitudinal testing in a consistent manner. In the case involving appetite, while we recognize that question 21 from the GDS-Long Form does assess appetite, we also found question 5 from the Montgomery and Åsberg (MADRS) Depression Rating Scale that may test the appetite subdomain to a higher degree of specificity. In this instance, we elected to include the MADRS based on our previous work that the metric can be utilized to measure an individual’s appetite.^8^

Stroke recovery is a dynamic process, and we have shown that mean perception scores express different recovery patterns over time based on the metric being used. Accordingly, it is also important to recognize that the subdomains of perception can vary in how long they take to recover. For example, some patients may experience a brief loss of consciousness from a stroke, whereas other aspects of perception, such as sleep disturbances, appetite changes, or pain, can persist long-term.^59,60^ Therefore, this proposed assessment battery can serve as a useful guide to measure perception recovery systematically, allowing for more precise monitoring of patient progress in a variety of critical components of perception. However, complete testing using the full metrics remains essential in certain contexts, especially if clinicians want a general measure of perception. To the best of our knowledge, there is currently no validated way to combine these individual items from our assessment battery together to form a suitable scoring system. Thus, while targeted assessments, such as the one we have proposed, provide flexibility, they function more as a guide to assessing stroke recovery and do not replace the need for comprehensive evaluations.

This review provides novel insights into the evolution of perception-based recovery following stroke, through the lens of a variety of metrics testing its relevant subdomains. With the large number of studies included using a comprehensive, structured search, alongside a substantial sample of patients, this study aims to characterize mean levels of perception recovery in standard stroke populations, allowing clinicians to compare patients’ recovery trajectories with the expected change in the literature.

## Limitations

This study is somewhat limited by its inclusion criteria, particularly through the requirement of all studies to be published in English, potentially leading to the exclusion of valuable data in standard stroke populations originating from non-English speaking regions. We also excluded studies with patients who had prior neurological disability or interventional treatment beyond the standard of care, which may prevent the generalizability of our findings to these populations. Another key limitation is the usage of iterative and/or mixed modeling in certain metrics, which at times, can fit recovery trajectories beyond a realistic level. While we prevented excessive fitting models as much as possible, we used polynomial models in one instance due to iterative modeling showing a small but significant decrease in scores following increasing logarithmic growth, which was unable to be modeled otherwise using standard patterns of growth. Modeling itself is inherently limited by its approximations of highly unpredictable and fluctuating patterns to predictable trajectories, especially for metrics with shorter follow-up periods where it is possible inaccurate models may be the most predictive. We also did not separate recovery in patients with ischemic vs. hemorrhagic stroke, with potential differences in the trajectory of perception function over time across cohorts. Finally, our table outlining the specific items in each metric corresponding to the relevant subdomain, as of now, cannot be used as a combined assessment battery for the domain of perception, due to vastly different scoring systems that cannot be combined. Instead, it solely serves as a guide for clinicians to specifically measure subdomain-specific recovery when required, with further validation required for broader applications.

The data we obtained is also somewhat limited by smaller sample sizes and/or the number of studies, especially for metrics such as the BBS and WHOQOL-BREF with relatively small samples and shorter intervals of recovery following stroke. Moreover, we had fewer studies reporting longitudinal scores at larger follow-up intervals across metrics, leading to higher CV and potentially inaccurate trajectories, such as those seen up to 10 years in the FMA-UE. The majority of our studies were also reported from centers in high-income countries, and hence may not be representative of standard recovery in populations in middle and low-income countries, alongside those with substantial differences in the standard of care. The recovery trajectories we described also only show the evolution of mean scores (±95% CI) of the metrics in a standard population over time, and do not provide information on where a patient’s score should be in comparison to the population mean. Due to differences in thresholds and a scarcity of studies depicting the expected percentile of scores in a cohort of stroke patients, further work is required before these results can be used to directly compare patient recovery.

## Conclusions

This study demonstrates the trajectory of stroke recovery within the domain of perception, particularly through the metrics of the NIHSS, EQ-5D, FMA, MMSE, BBS, GDS, and WHOQOL-BREF to ensure the inclusion of all subdomains. Through iterative modeling, we characterize the expected longitudinal changes in scores across metrics in standard stroke populations, alongside highlighting the relevant items of each metric corresponding to each subdomain of perception. Through these findings, we hope this study can guide clinicians in better understanding the evolution of perception-based physiology in stroke patients, alongside encouraging further research exploring the validity of our findings and their application to identifying the prognosis of stroke recovery in individuals.

## Data Availability

All data requests can be raised with the corresponding author.

## Contributions

Conceptualization, Y.A. and C.P.K.; methodology, Y.A., R.A., M.L., K.N. and C.P.K.; formal analysis, Y.A., K.N. and C.P.K.; investigation, Y.A., R.A., M.L., K.N. and C.P.K.; data curation, Y.A., R.A., M.L., K.N. and C.P.K.; writing—original draft preparation, Y.A. and K.N..; writing—review and editing, Y.A., K.N., C.P.K; supervision, C.P.K.; project administration, C.P.K. All authors have read and agreed to the published version of the manuscript.

## Sources of Funding

This study did not require or use funding.

## Disclosures

The authors have nothing to disclose.

## Data Availability

All data requests can be raised with the corresponding author.

## Statement of Ethics

A statement of ethics is not applicable because this study is based exclusively on published literature.

## Notes

### Competing Interest Statement

The authors have declared no competing interest.

### Author Declarations

An IRB/oversight body was not needed for this study.

## References

1. Imoisili OE, Chung A, Tong X, Hayes DK, Loustalot F. Prevalence of Stroke - Behavioral Risk Factor Surveillance System, United States, 2011-2022. MMWR Morb Mortal Wkly Rep. 2024;73:449–455. doi: 10.15585/mmwr.mm7320a1

2. Feigin VL, Brainin M, Norrving B, Martins S, Sacco RL, Hacke W, Fisher M, Pandian J, Lindsay P. World Stroke Organization (WSO): Global Stroke Fact Sheet 2022. Int J Stroke. 2022;17:18–29. doi: 10.1177/17474930211065917

3. Katan M, Luft A. Global Burden of Stroke. Semin Neurol. 2018;38:208–211. doi: 10.1055/s-0038-1649503

4. Kim YW. Update on Stroke Rehabilitation in Motor Impairment. Brain Neurorehabil. 2022;15:e12. doi: 10.12786/bn.2022.15.e12

5. Avan A, Digaleh H, Di Napoli M, Stranges S, Behrouz R, Shojaeianbabaei G, Amiri A, Tabrizi R, Mokhber N, Spence JD, et al. Socioeconomic status and stroke incidence, prevalence, mortality, and worldwide burden: an ecological analysis from the Global Burden of Disease Study 2017. BMC Med. 2019;17:191. doi: 10.1186/s12916-019-1397-3

6. Brady MC, Kelly H, Godwin J, Enderby P, Campbell P. Speech and language therapy for aphasia following stroke. Cochrane Database Syst Rev. 2016;2016:CD000425. doi: 10.1002/14651858.CD000425.pub4

7. Vanhook P. The domains of stroke recovery: a synopsis of the literature. J Neurosci Nurs. 2009;41:6–17. doi: 10.1097/jnn.0b013e31819345e4

8. Akkara Y, Afreen R, Lemonick M, Paz SG, Rifi Z, Tosto J, Putrino D, Mocco J, Bederson J, Dangayach N, et al. Standardizing Domains and Metrics of Stroke Recovery: A Systematic Review. Brain Sci. 2024;14. doi: 10.3390/brainsci14121267

9. Hazelton C, McGill K, Campbell P, Todhunter-Brown A, Thomson K, Nicolson DJ, Cheyne JD, Chung C, Dorris L, Gillespie DC, et al. Perceptual Disorders After Stroke: A Scoping Review of Interventions. Stroke. 2022;53:1772–1787. doi: 10.1161/STROKEAHA.121.035671

10. Elkholi SMA, Abdelwahab MK, Abdelhafeez M. Impact of the smell loss on the quality of life and adopted coping strategies in COVID-19 patients. Eur Arch Otorhinolaryngol. 2021;278:3307–3314. doi: 10.1007/s00405-020-06575-7

11. Edmans J, Towle D, Lincoln N. The recovery of perceptual problems after stroke and the impact on daily life. Clinical Rehabilitation. 1991;5:301–309. doi: 10.1177/026921559100500406

12. Colwell MJ, Demeyere N, Vancleef K. Visual perceptual deficit screening in stroke survivors: evaluation of current practice in the United Kingdom and Republic of Ireland. Disabil Rehabil. 2022;44:6620–6632. doi: 10.1080/09638288.2021.1970246

13. Dutta TM, Josiah AF, Cronin CA, Wittenberg GF, Cole JW. Altered taste and stroke: a case report and literature review. Top Stroke Rehabil. 2013;20:78–86. doi: 10.1310/tsr2001-78

14. Rowe FJ, Walker M, Rockliffe J, Pollock A, Noonan C, Howard C, Glendinning R, Feechan R, Currie J. Care provision for poststroke visual impairment. J Stroke Cerebrovasc Dis. 2015;24:1131–1144. doi: 10.1016/j.jstrokecerebrovasdis.2014.12.035

15. Ballester BR, Maier M, Duff A, Cameirao M, Bermudez S, Duarte E, Cuxart A, Rodriguez S, San Segundo Mozo RM, Verschure P. A critical time window for recovery extends beyond one-year post-stroke. J Neurophysiol. 2019;122:350–357. doi: 10.1152/jn.00762.2018

16. Wade DT, Wood VA, Hewer RL. Recovery after stroke--the first 3 months. J Neurol Neurosurg Psychiatry. 1985;48:7–13. doi: 10.1136/jnnp.48.1.7

17. Dros J, Segiet N, Poczatek G, Klimkowicz-Mrowiec A. Five-year stroke prognosis. Influence of post-stroke delirium and post-stroke dementia on mortality and disability (Research Study - Part of the PROPOLIS Study). Neurol Sci. 2024;45:1109–1119. doi: 10.1007/s10072-023-07129-5

18. Adoukonou T, Agbetou M, Bangbotche R, Kossi O, Fotso Mefo P, Magne J, Houinato D, Preux PM, Lacroix P. Long-Term Mortality of Stroke Survivors in Parakou: 5-Year Follow-Up. J Stroke Cerebrovasc Dis. 2020;29:104785. doi: 10.1016/j.jstrokecerebrovasdis.2020.104785

19. Hart CL, Hole DJ, Smith GD. Comparison of risk factors for stroke incidence and stroke mortality in 20 years of follow-up in men and women in the Renfrew/Paisley Study in Scotland. Stroke. 2000;31:1893–1896. doi: 10.1161/01.str.31.8.1893

20. Fugl-Meyer AR, Jaasko L, Leyman I, Olsson S, Steglind S. The post-stroke hemiplegic patient. 1. a method for evaluation of physical performance. Scand J Rehabil Med. 1975;7:13–31.

21. Kwakkel G, Kollen B, Twisk J. Impact of time on improvement of outcome after stroke. Stroke. 2006;37:2348–2353. doi: 10.1161/01.STR.0000238594.91938.1e

22. Hordacre B, Austin D, Brown KE, Graetz L, Parees I, De Trane S, Vallence AM, Koblar S, Kleinig T, McDonnell MN, et al. Evidence for a Window of Enhanced Plasticity in the Human Motor Cortex Following Ischemic Stroke. Neurorehabil Neural Repair. 2021;35:307–320. doi: 10.1177/1545968321992330

23. Zeiler SR, Krakauer JW. The interaction between training and plasticity in the poststroke brain. Curr Opin Neurol. 2013;26:609–616. doi: 10.1097/WCO.0000000000000025

24. Prabhakaran S, Zarahn E, Riley C, Speizer A, Chong JY, Lazar RM, Marshall RS, Krakauer JW. Inter-individual variability in the capacity for motor recovery after ischemic stroke. Neurorehabil Neural Repair. 2008;22:64–71. doi: 10.1177/1545968307305302

25. Ingwersen T, Wolf S, Birke G, Schlemm E, Bartling C, Bender G, Meyer A, Nolte A, Ottes K, Pade O, et al. Long-term recovery of upper limb motor function and self-reported health: results from a multicenter observational study 1 year after discharge from rehabilitation. Neurol Res Pract. 2021;3:66. doi: 10.1186/s42466-021-00164-7

26. Balestroni G, Bertolotti G. [EuroQol-5D (EQ-5D): an instrument for measuring quality of life]. Monaldi Arch Chest Dis. 2012;78:155–159. doi: 10.4081/monaldi.2012.121

27. Luengo-Fernandez R, Gray AM, Bull L, Welch S, Cuthbertson F, Rothwell PM, Oxford Vascular S. Quality of life after TIA and stroke: ten-year results of the Oxford Vascular Study. Neurology. 2013;81:1588–1595. doi: 10.1212/WNL.0b013e3182a9f45f

28. Rabin R, de Charro F. EQ-5D: a measure of health status from the EuroQol Group. Ann Med. 2001;33:337–343. doi: 10.3109/07853890109002087

29. Bushnell CD, Reeves MJ, Zhao X, Pan W, Prvu-Bettger J, Zimmer L, Olson D, Peterson E. Sex differences in quality of life after ischemic stroke. Neurology. 2014;82:922–931. doi: 10.1212/WNL.0000000000000208

30. Kurlowicz L, Wallace M. The Mini-Mental State Examination (MMSE). J Gerontol Nurs. 1999;25:8–9. doi: 10.3928/0098-9134-19990501-08

31. Kalaria RN, Akinyemi R, Ihara M. Stroke injury, cognitive impairment and vascular dementia. Biochim Biophys Acta. 2016;1862:915–925. doi: 10.1016/j.bbadis.2016.01.015

32. Rost NS, Brodtmann A, Pase MP, van Veluw SJ, Biffi A, Duering M, Hinman JD, Dichgans M. Post-Stroke Cognitive Impairment and Dementia. Circ Res. 2022;130:1252-1271. doi: 10.1161/CIRCRESAHA.122.319951

33. Greenberg SM, Ziai WC, Cordonnier C, Dowlatshahi D, Francis B, Goldstein JN, Hemphill JC, 3rd, Johnson R, Keigher KM, Mack WJ, et al. 2022 Guideline for the Management of Patients With Spontaneous Intracerebral Hemorrhage: A Guideline From the American Heart Association/American Stroke Association. Stroke. 2022;53:e282–e361. doi: 10.1161/STR.0000000000000407

34. Brodtmann A, Werden E, Khlif MS, Bird LJ, Egorova N, Veldsman M, Pardoe H, Jackson G, Bradshaw J, Darby D, et al. Neurodegeneration Over 3 Years Following Ischaemic Stroke: Findings From the Cognition and Neocortical Volume After Stroke Study. Front Neurol. 2021;12:754204. doi: 10.3389/fneur.2021.754204

35. Brunelli S, Giannella E, Bizzaglia M, De Angelis D, Sancesario GM. Secondary neurodegeneration following Stroke: what can blood biomarkers tell us? Front Neurol. 2023;14:1198216. doi: 10.3389/fneur.2023.1198216

36. Kwah LK, Diong J. National Institutes of Health Stroke Scale (NIHSS). J Physiother. 2014;60:61. doi: 10.1016/j.jphys.2013.12.012

37. Berg K, Wood-Dauphine S, Williams JI, Gayton D. Measuring balance in the elderly: preliminary development of an instrument. Physiotherapy Canada. 1989;41:304–311. doi: 10.3138/ptc.41.6.304

38. Buvarp D, Rafsten L, Abzhandadze T, Sunnerhagen KS. A cohort study on longitudinal changes in postural balance during the first year after stroke. BMC Neurol. 2022;22:324. doi: 10.1186/s12883-022-02851-7

39. Pahlman U, Gutierrez-Perez C, Savborg M, Knopp E, Tarkowski E. Cognitive function and improvement of balance after stroke in elderly people: the Gothenburg cognitive stroke study in the elderly. Disabil Rehabil. 2011;33:1952–1962. doi: 10.3109/09638288.2011.553703

40. Divandari N, Bird ML, Vakili M, Jaberzadeh S. The Association Between Cognitive Domains and Postural Balance among Healthy Older Adults: A Systematic Review of Literature and Meta-Analysis. Curr Neurol Neurosci Rep. 2023;23:681–693. doi: 10.1007/s11910-023-01305-y

41. Yu HX, Wang ZX, Liu CB, Dai P, Lan Y, Xu GQ. Effect of Cognitive Function on Balance and Posture Control after Stroke. Neural Plast. 2021;2021:6636999. doi: 10.1155/2021/6636999

42. Turunen KEA, Laari SPK, Kauranen TV, Uimonen J, Mustanoja S, Tatlisumak T, Poutiainen E. Domain-Specific Cognitive Recovery after First-Ever Stroke: A 2-Year Follow-Up. J Int Neuropsychol Soc. 2018;24:117–127. doi: 10.1017/S1355617717000728

43. Lee EY, Na Y, Cho M, Hwang YM, Noh JS, Kwon HK, Pyun SB. Clinical Factors Associated With Balance Function in the Early Subacute Phase After Stroke. Am J Phys Med Rehabil. 2022;101:203–210. doi: 10.1097/PHM.0000000000001856

44. Purohit R, Wang S, Dusane S, Bhatt T. Age-related differences in reactive balance control and fall-risk in people with chronic stroke. Gait Posture. 2023;102:186–192. doi: 10.1016/j.gaitpost.2023.03.011

45. Volz M, Mobus J, Letsch C, Werheid K. The influence of early depressive symptoms, social support and decreasing self-efficacy on depression 6 months post-stroke. J Affect Disord. 2016;206:252–255. doi: 10.1016/j.jad.2016.07.041

46. Yesavage JA, Brink TL, Rose TL, Lum O, Huang V, Adey M, Leirer VO. Development and validation of a geriatric depression screening scale: a preliminary report. J Psychiatr Res. 1982;17:37–49. doi: 10.1016/0022-3956(82)90033-4

47. Dong L, Williams LS, Brown DL, Case E, Morgenstern LB, Lisabeth LD. Prevalence and Course of Depression During the First Year After Mild to Moderate Stroke. J Am Heart Assoc. 2021;10:e020494. doi: 10.1161/JAHA.120.020494

48. Liu L, Xu M, Marshall IJ, Wolfe CD, Wang Y, O’Connell MD. Prevalence and natural history of depression after stroke: A systematic review and meta-analysis of observational studies. PLoS Med. 2023;20:e1004200. doi: 10.1371/journal.pmed.1004200

49. Wong FY, Yang L, Yuen JWM, Chang KKP, Wong FKY. Assessing quality of life using WHOQOL-BREF: a cross-sectional study on the association between quality of life and neighborhood environmental satisfaction, and the mediating effect of health-related behaviors. BMC Public Health. 2018;18:1113. doi: 10.1186/s12889-018-5942-3

50. Vahedi S. World Health Organization Quality-of-Life Scale (WHOQOL-BREF): Analyses of Their Item Response Theory Properties Based on the Graded Responses Model. Iran J Psychiatry. 2010;5:140–153.

51. Horgan NF, O’Regan M, Cunningham CJ, Finn AM. Recovery after stroke: a 1-year profile. Disabil Rehabil. 2009;31:831–839. doi: 10.1080/09638280802355072

52. Kainz A, Meisinger C, Linseisen J, Kirchberger I, Zickler P, Naumann M, Ertl M. Changes of Health-Related Quality of Life Within the 1st Year After Stroke-Results From a Prospective Stroke Cohort Study. Front Neurol. 2021;12:715313. doi: 10.3389/fneur.2021.715313

53. Marten O, Greiner W. Exploring differences and similarities of EQ-5D-3L, EQ-5D-5L and WHOQOL-OLD in recipients of aged care services in Germany. PLoS One. 2023;18:e0290606. doi: 10.1371/journal.pone.0290606

54. Skevington SM, Lotfy M, O’Connell KA, Group W. The World Health Organization’s WHOQOL-BREF quality of life assessment: psychometric properties and results of the international field trial. A report from the WHOQOL group. Qual Life Res. 2004;13:299–310. doi: 10.1023/B:QURE.0000018486.91360.00

55. Pedersen SG, Friborg O, Heiberg GA, Arntzen C, Stabel HH, Thrane G, Nielsen JF, Anke A. Stroke-Specific Quality of Life one-year post-stroke in two Scandinavian country-regions with different organisation of rehabilitation services: a prospective study. Disabil Rehabil. 2021;43:3810–3820. doi: 10.1080/09638288.2020.1753830

56. Pucciarelli G, Brugnera A, Greco A, Petrizzo A, Simeone S, Vellone E, Alvaro R. Stroke disease-specific quality of life trajectories after rehabilitation discharge and their sociodemographic and clinical associations: A longitudinal, multicentre study. J Adv Nurs. 2021;77:1856–1866. doi: 10.1111/jan.14722

57. Crichton SL, Bray BD, McKevitt C, Rudd AG, Wolfe CD. Patient outcomes up to 15 years after stroke: survival, disability, quality of life, cognition and mental health. J Neurol Neurosurg Psychiatry. 2016;87:1091–1098. doi: 10.1136/jnnp-2016-313361

58. Piscicelli C, Perennou D. Visual verticality perception after stroke: A systematic review of methodological approaches and suggestions for standardization. Ann Phys Rehabil Med. 2017;60:208–216. doi: 10.1016/j.rehab.2016.02.004

59. Ali M, Tibble H, Brady MC, Quinn TJ, Sunnerhagen KS, Venketasubramanian N, Shuaib A, Pandyan A, Mead G, Collaboration V. Prevalence, Trajectory, and Predictors of Poststroke Pain: Retrospective Analysis of Pooled Clinical Trial Data Set. Stroke. 2023;54:3107–3116. doi: 10.1161/STROKEAHA.123.043355

60. Duss SB, Bauer-Gambelli SA, Bernasconi C, Dekkers MPJ, Gorban-Peric C, Kuen D, Seiler A, Oberholzer M, Alexiev F, Lippert J, et al. Frequency and evolution of sleep-wake disturbances after ischemic stroke: A 2-year prospective study of 437 patients. Sleep Med. 2023;101:244–251. doi: 10.1016/j.sleep.2022.10.007

